# The Maine Vein of Marshall Ethanol Experience: Learning Curve, Efficacy, and Safety

**DOI:** 10.1101/2022.08.08.22278568

**Authors:** Jordan S Leyton-Mange, Kunal Tandon, Edward Y Sze, Charles M Carpenter, Henry W Sesselberg

**Author notes:** **Contact Information:**, Correspondence: Jordan Leyton-Mange, MaineHealth Cardiology, 96 Campus Drive, Scarborough, ME 04074. **Author contributions:** All authors contributed to the study conception and design. J.L.M was the primary operator for the procedures described and performed the data collection, analysis, and preparation of the first draft of the manuscript. All authors commented on subsequent versions as well as read and approved the final manuscript. **Compliance with Ethical Standards**. Ethical approval:* This study was approved by the Maine Medical Center Institutional Review Board and performed in accordance with the Declaration of Helsinki. Informed consent:* This is a retrospective study performed by chart review and as such, informed consent was waved by the Institutional Review Board.

## Abstract

**Background:** The marginal benefit of ethanol infusion into the VOM as an adjunct to atrial fibrillation ablation has shown promise in a single randomized study and case series from very experienced centers. However, adoption has not been widespread and the impact on real-world outcomes outside of leading centers is not established.

**Objective:** To understand the learning curve, and explore procedural outcomes and safety with VOM ethanol infusion from a large single medical center.

**Methods:** All atrial ablation cases wherein VOM ethanol infusion was attempted were identified from the time of the program’s inception in 2019 at Maine Medical Center (Portland, ME). Our technical approach, procedural success and efficacy rates, and complications were adjudicated from the medical record.

**Results:** The overall VOM ethanol infusion success was 90%. Infusion success rates improved and fluoroscopy utilization decreased with experience. Arrhythmia recurrence was 86% after a mean follow-up of 9.5 months, with an arrhythmia-freedom probability of 80% at 12 months. Complications occurred in 5.4% of patients, including a 3.1% risk of delayed tamponade.

**Conclusion:** In our single center experience, VOM ethanol infusion was feasible with a high technical success rate and excellent arrhythmia freedom in follow-up. These positive results are balanced against a concerning rate of delayed tamponade.

## Introduction

Since the discovery of pulmonary venous triggers,^1^ catheter ablation with pulmonary vein isolation (PVI) has matured as an important therapy for patients with atrial fibrillation (AF). However despite advances in ablation technology, for non-paroxysmal forms of AF, outcomes remain suboptimal with PVI alone.^2, 3^ Adjunctive strategies addressing the atrial substrate or targeting of non-pulmonary vein triggers have not shown consistent efficacy across studies and may carry additional complication risks or higher rates of recurrent organized atrial tachycardias. Recently, ethanol infusion into the vein of Marshall (VOM) has gained popularity with the publication of the VENUS randomized trial^4^ and the positive accumulating experience reported from a single center in Bordeaux.^5, 6^ However, adoption has been tempered due to the technical requirements of the procedure and unclear dominant mechanism of putative benefit. Thus, more widespread experience is needed to clarify the real-world impact on procedural success. This report aims to describe a single center experience with VOM ethanol infusion over three years, with description of technical approach, associated procedural success rates, and complications.

## Material and Methods

### Study population

We retrospectively analyzed all atrial ablation cases wherein VOM ethanol infusion was attempted between August of 2019 and May of 2022 at Maine Medical Center (Portland, ME). This study was approved by the Institutional Review Board and performed in accordance with the Declaration of Helsinki.

### Catheter ablation

Ablations were performed under general anesthesia with low tidal volume ventilation and uninterrupted anticoagulation. Three dimensional guidance was performed with the CARTO 3 system (Biosense Webster). Heparin infusion was utilized to maintain an activated clotting time of 350 seconds. Pacing from either the coronary sinus (CS) or right ventricle was performed to reduce the stroke volume and variability in cardiac motion. All procedures utilized irrigated radiofrequency with the STSF catheter aiming for interlesion distances of 4mm (Biosense Webster). In addition to VOM ethanol infusion, the standard lesion set for patients with persistent AF in this study consisted of PVI, left atrial posterior wall isolation (PWI), CS isolation, posterior mitral isthmus line (MIL), and a cavotricuspid isthmus line (CTI) (Figure 1). The mitral isthmus was ablated with air-filled balloon occlusion of the CS in almost all cases to improve lesion transmurality near the annulus.^7^ In the subset of patients both with persistent and occasionally paroxysmal AF wherein VOM ethanol was not preplanned and rather performed *ad hoc* (most commonly for induced mitral annular flutter in patients with myopathic low voltage regions, Table 1), MIL, CS isolation, and PWI was still systematically performed. The sought end points for CS isolation and PWI included elimination of local electrograms and lack of pace capture of the atrium with high output bipolar pacing from within the isolated structure (20mA, 2msec). The proximal third of the CS was not ablated to avoid iatrogenic atrioventricular (AV) block. For the first half of the enrollment period, luminal temperature monitoring was used for esophageal protection and occasionally, PWI was avoided in favor of a reinforced roof line if the esophageal risk was deemed too great because of esophageal heating or close proximity to the posterior wall on intracardiac echo (ICE). Towards the latter half of the study, patients underwent active esophageal cooling at 4°C (Enso ETM, Attune Medical, Chicago, IL). Rarely, CTI ablation was avoided if typical atrial flutter was not inducible and the appearance of the region on ICE appeared anatomically challenging. Adenosine was used to assess for dormant conduction and in select procedures, isoproterenol infusion was used to disclose non-PV foci.

**Figure 1.**
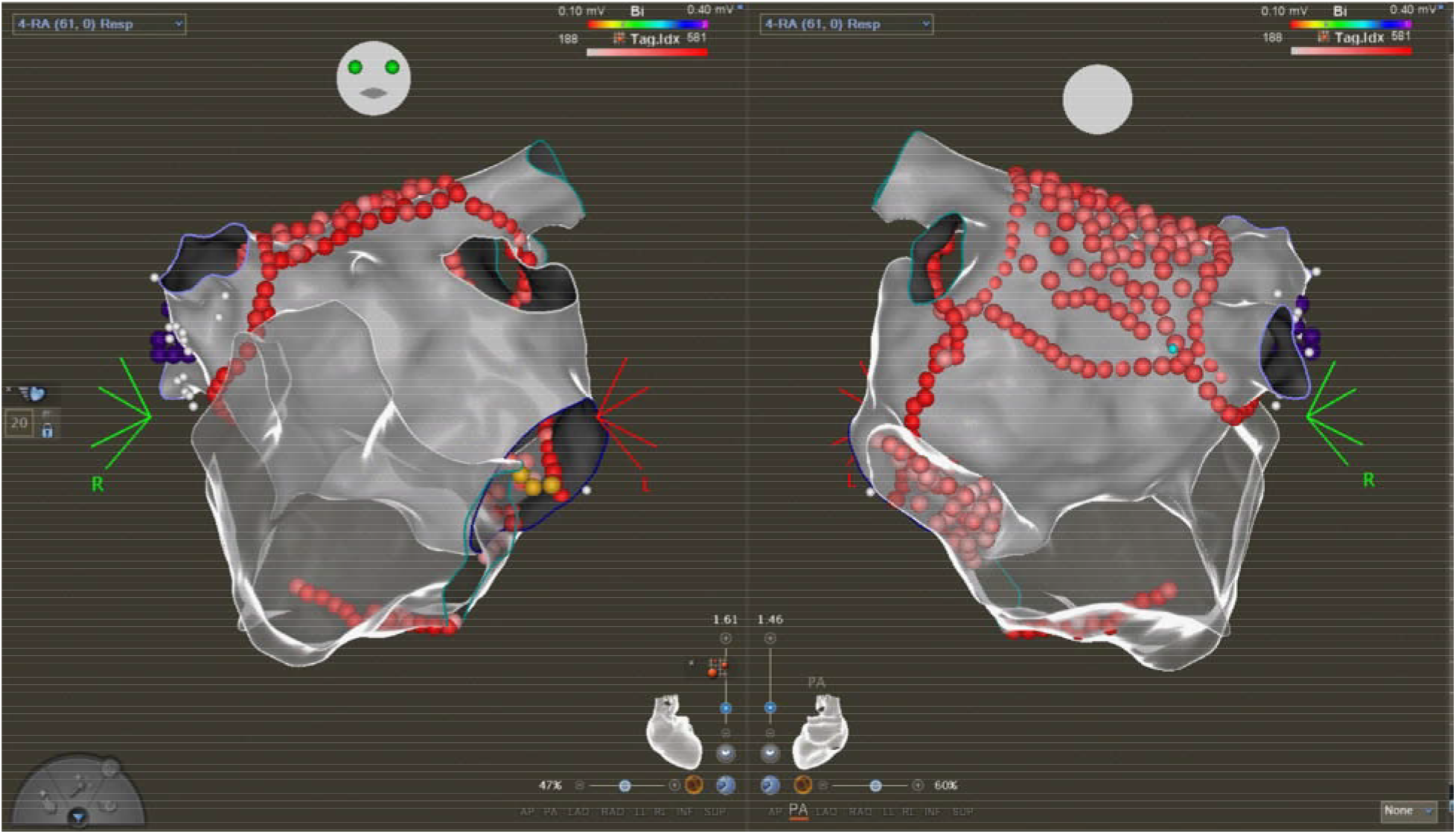
Three dimensional maps depicting the standard lesion set used for patients in this study. In addition to VOM ethanol infusion, PVI, PWI, CS isolation, MIL and CTI lines were systematically performed.

**Table 1.** Patient characteristics and procedural details. Abbreviations: pAF – paroxysmal AF, persAF – persistent AF, LSPAF – long-standing persistent AF, AVNRT – AV nodal reentrant tachycardia.

|  |  |
| --- | --- |
| Total patients | 129 |
| *Paroxysmal, Persistent, Long-Standing Persistent* | *(23 pAF, 90 persAF, 16 LSPAF)* |
| Average age (years) | 64.3 |
| Male | 67.4% |
| Left atrial diameter, avg (mm) | 44.3 |
| Cases by year |  |
| 2019 | 5 |
| 2020 | 20 |
| 2021 | 71 |
| 2022 | 33 |
| Index AF ablations | 90 |
| *Preplanned VOM* | *74% (67: 1 pAF, 52 persAF, 14 LSPAF)* |
| *Ad hoc VOM* | *26% (23: 14 pAF, 9 persAF)* |
| *Standard lesion set only* | *88% (79)* |
| *SVC ablation/isolation* | *5.6% (5)* |
| *Additional arrhythmias (AVNRT, atypical flutter)* | *3.3% (3)* |
| *Non-PV triggers disclosed and ablated* | *1.1% (1)* |
| *Extension of PWI to CS* | *2.2% (2)* |
| Redo Procedures | 39 |
| 1 prior; 2 prior; 3 prior | 32; 5; 2 |
| *Preplanned VOM* | *95% (37: 7 pAF, 28 persAF, 2 LSPAF)* |
| *Ad hoc VOM* | *5.1% (2: 1 pAF, 1 persAF)* |
| *Permanent PVI upon initial mapping* | *56% (22)* |
| *Standard lesion set only* | *23% (9)* |
| *SVC ablation/isolation* | *69% (27)* |
| *Additional arrhythmias (AVNRT, atypical flutter)* | *18% (7)* |
| *Non-PV triggers disclosed and ablated* | *5.1% (2)* |
| *Extension of PWI to CS* | *7.6% (3)* |
| VOM ethanol infusion success | 90% |
| 2019-2020 success | 76% |
| 2022 success | 100% |
| Reasons for VOM Ethanol failure | 9.3% (12) |
| *CS could not be cannulated* | *1* |
| *Left persistent SVC* | *1* |
| *Wire perforation* | *1* |
| *Incorrect vein targeted* | *2* |
| *VOM could not be cannulated* | *3* |
| *VOM not identified* | *4* |
| VOM Dissection | 8.5% |
| VOM Perforation | 1.6% |
| Fluoroscopy, avg (min) | 18.8 |
| 2019-2020 | 34.6 |
| 2022 | 10.5 |

### VOM ethanol infusion

VOM ethanol infusion was performed prior to transseptal puncture and systemic heparinization in the first several cases but following left atrial mapping in the subsequent ones. From a femoral approach, the CS was cannulated with an SL-2 sheath over a deflectable decapolar catheter. A floppy tip straight 0.35” guidewire (MagicTorque, Boston Scientific or Wholey wire, Medtronic) was advanced past the Vieussens valve into the great cardiac vein to retain access and a venogram was performed through the sheath in the right anterior oblique projection to highlight the VOM. Often the VOM was not identifiable on venography but was subsequently found either proximal to the sheath tip or just proximal to Vieussens valve. An IMA guide catheter was carefully advanced over another 0.35” guidewire into the CS. The guidewire was then exchanged for an 0.14” angioplasty wire preloaded with an over the wire balloon to occlude the proximal VOM (1.5mm-2.5mm range though typically 2mm, Emerge Dilation catheter, Boston Scientific). An 0.14” PT Graphix wire (Straight tip, 300cm, Boston Scientific) was used for the initial cases but all subsequent ones were performed with the low tip weight Suoh wire (0.3 gf, Asahi) to virtually eliminate the risk of wire perforation. After VOM ethanol infusion, CS ablation was systematically performed through the SL-2 sheath. A rare subset of cases were performed from a superior approach, including the initial case in the series as well as subsequent ones wherein the VOM could not be stably cannulated from a femoral approach. After proximal VOM occlusion, a selective venogram was performed and up to 4 injections of anhydrous ethanol were performed of 1-3 cc, each over 30-60 seconds. Most infections were approximately 2.5 cc (two 5 cc vials split across four injections). To reduce risk towards the end of the study after four cases of delayed pericardial effusions were noted, the infusion volume was decreased to four injections of 1-1.5 cc, depending on the size of the VOM. Echogenicity was usually noted on ICE and a low voltage zone of varying size and confluency was demonstrated on electroanatomical mapping. VOM venograms were not systemically performed between ethanol injections, fearing additive risk of dissection of the VOM from viscous iodinated contrast unless there was suspicion of poor occlusion, balloon movement, or dissection, particularly in the absence of a significant ethanol response. Due to a series of dissections noted on initial venography and the delayed pericardial effusions noted in this study, for the final twenty one cases in the series, systematic use of the initial VOM venogram was abandoned unless prompted by concern of poor ethanol delivery on imaging or mapping due to the perceived dissection risks from brisk injections of contrast.

### Outcomes

Patients were treated and followed according to local standards of care. Recurrences were adjudicated by the study authors based upon rhythm tracings in the medical record. Tracings from standard electrocardiography, wearable rhythm monitoring, and recordings from cardiac implantable electronic devices (CIED), whether scheduled or symptom triggered, were permitted and assessed. Recurrence was defined as recurrent atrial fibrillation, atrial flutter, or atrial tachycardia of at least 30 seconds duration, after at least 90 days post procedure (blanking period). Complications were determined from the medical record. Tamponade was defined as pericardial effusion requiring pericardiocentesis.

### Statistics

Most outcomes are presented as raw percentages. Survival probabilities were computed with a product limit (Kaplan Meier) approach, censoring patients at the time of recurrence.

## Results

### Procedural details

A total of 129 patients underwent attempted VOM ethanol infusion during the study period (Table 1). The average age was 64.3 years and 67.4% were male. Ninety were initial procedures for AF and 39 were redo procedures. Of the latter group, 56% had permanent PVI upon initial intraprocedural mapping. Most VOM infusions were preplanned, but a substantial proportion, including 26% in the group of initial procedures, were *ad hoc*. All but one of these were performed a*d hoc* to facilitate block for induced mitral annular flutter, with the additional one done in the case of a suspected ligament of Marshall trigger for AF after PVI was completed. The *ad hoc* group encompassed nearly the entirety of patients with paroxysmal AF undergoing VOM infusion during an index procedure. Ablation was restricted to the standard lesion set as depicted in Figure 1 in 88% of the patients taken for an initial AF ablation but the majority of redo cases had additional lesions, most commonly SVC isolation/ablation (69%).

The success rate of VOM infusion was 90%, though improved in follow-up from a 76% success in the initial years of the study to 100% in 2022. Reasons for failure of ethanol delivery in twelve patients included failure to cannulate the CS (1), a left persistent superior vena cava (SVC) (1), a wire perforation with avoidance of ethanol delivery (1), targeting of the incorrect vein (2), failure to cannulate the VOM (3), and failure to identify the VOM (4). Total case fluoroscopy averaged 18.8 minutes but decreased during the study from 34.6 minutes during the first two years of the study to 10.5 minutes during 2022 (Figure 2). VOM perforation occurred in two patients (1.6%) including one wherein ethanol was not delivered. VOM dissection occurred in 8.5% of patients. These events and procedural success are depicted by procedure order in Figure 2A. All attempts at acute mitral isthmus block were successful, irrespective of whether the VOM ethanol infusion was completed.

**Figure 2.**
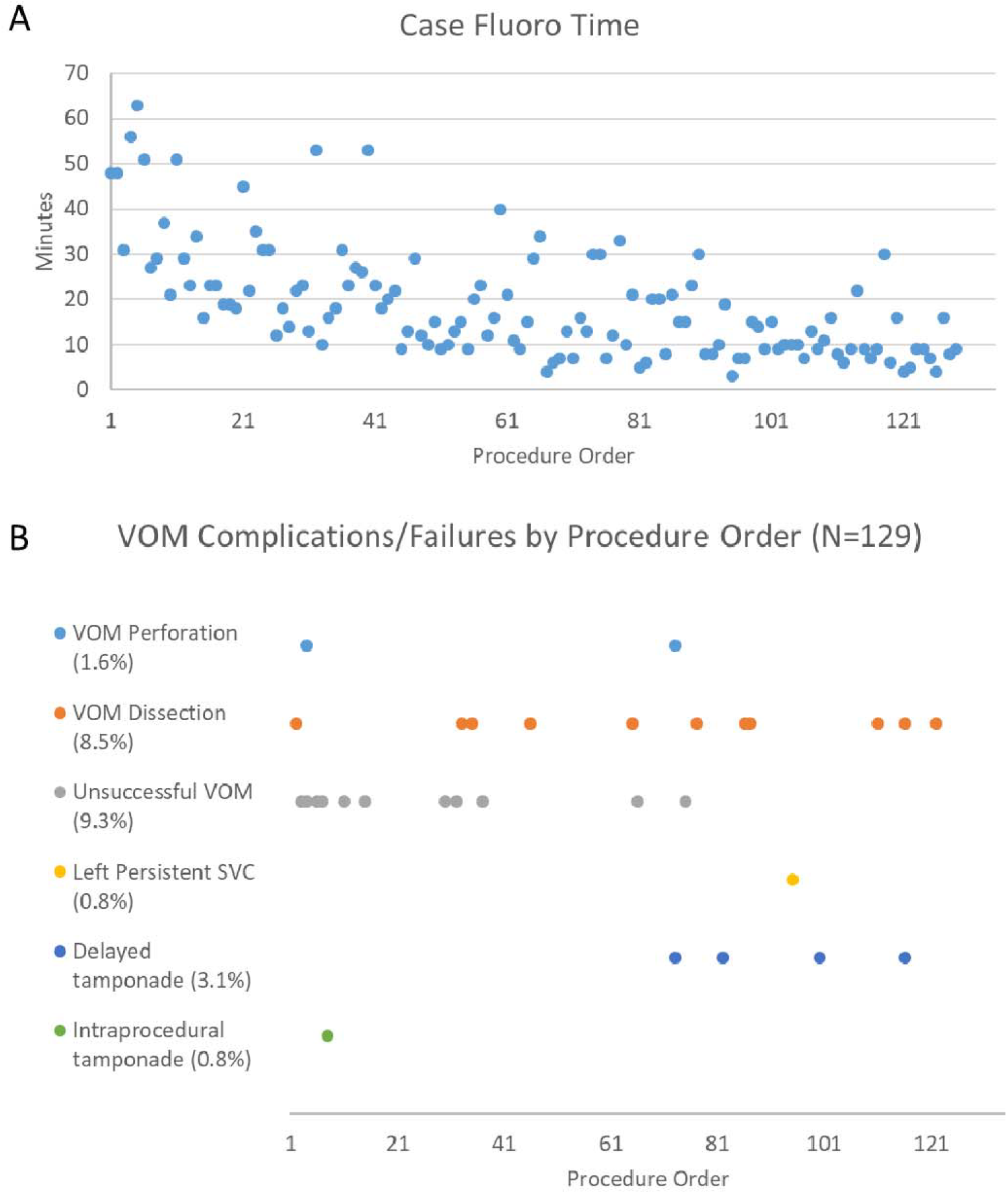
(A) Total case fluoroscopy time and (B) VOM ethanol infusion complications catalogued by procedure order (N=129).

### Efficacy

A total of 94 patients had at least one rhythm assessment at least 90 days post procedure and were included in analysis of efficacy outcomes. The follow-up duration ranged from 3.0 to 30.1 months with an average duration of 9.5 months. Rhythm assessments were made with CIEDs in 17% of patients, prescribed wearable monitors in 41.5%, and standard electrocardiography alone in 41.5% of patients. The overall recurrence rate was 14% after a 90 day blanking period. Six (6%) patients remained on antiarrhythmic drugs. Only five (5%) patients experienced clinical failure, a subjective assessment by the authors conveying failure of arrhythmia control (on or off antiarrhythmic drugs) or a need for further procedures. The majority of the others were modest and detailed in a Supplemental Table 1. Survival probabilities are shown in Figure 3, estimating an 80% arrhythmia free survival at twelve months following the initial VOM infusion ablation. The respective arrhythmia freedom probabilities at twelve months were 80% when restricting analysis to initial procedures (N=65 procedures), and 79% when only analyzing persistent AF patients undergoing an initial procedure (N=54 procedures).

**Figure 3.**
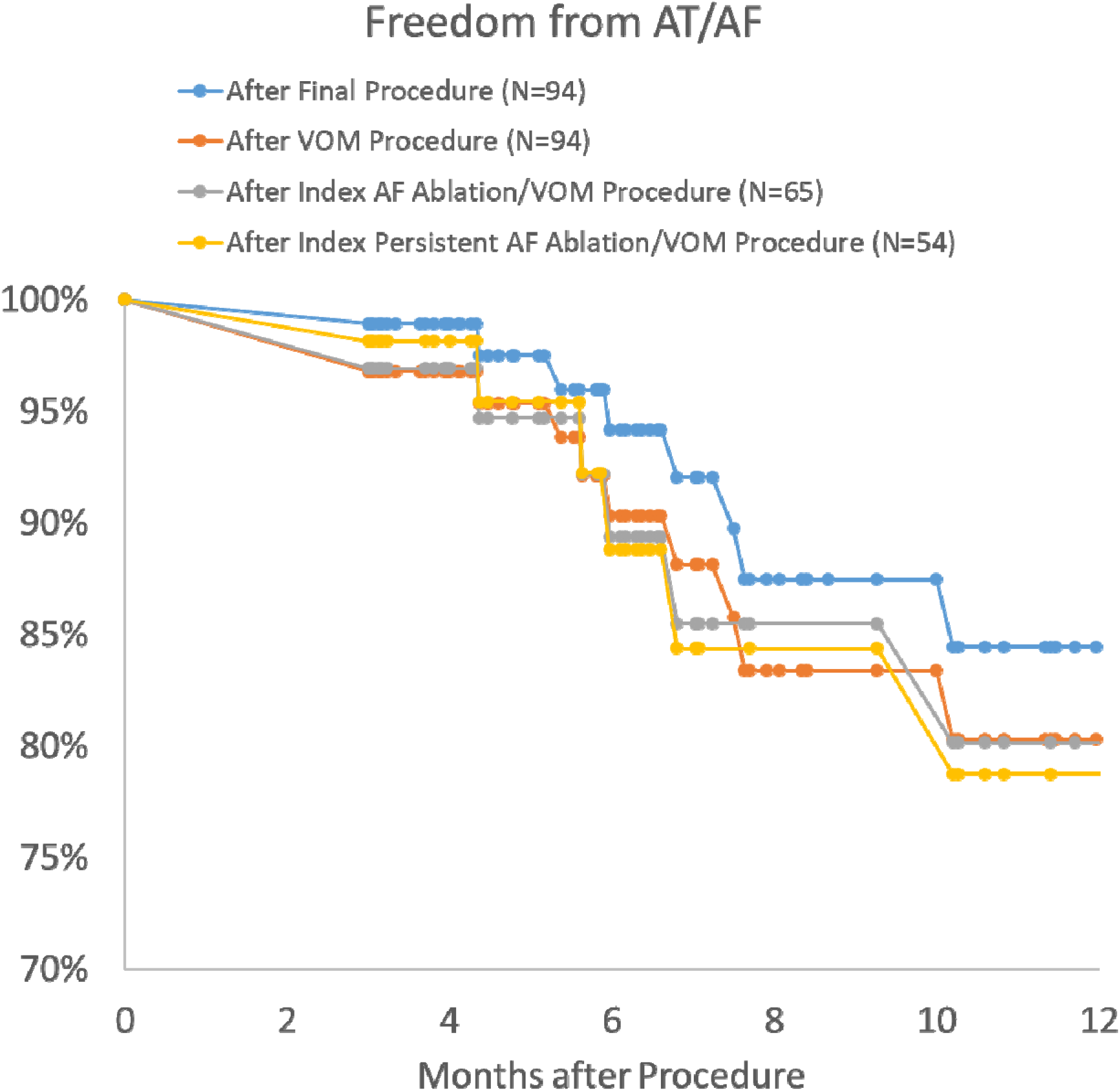
Kaplan Meier survival probabilities from atrial tachyarrhythmias for 12 months following ablation after the final ablation procedure including four patients that had redo ablations following the VOM procedure (blue, N=94), after the VOM procedure (N=94), after only initial AF ablations (N=65), and after only initial ablations performed for patients with persistent AF (N=54).

Four patients were taken for redo ablations after the VOM ethanol infusion procedure. Two of these patients experienced atypical flutter, both corresponding to circuits involving the posterior wall. The mitral isthmus required reablation in one of those patients though it was not mediating the recurrent arrhythmia. The other two patients each experienced recurrent paroxysmal AF. The first required early reablation for repeated cardioversions in emergency room settings with poorly controlled rates. In this case, the left veins, posterior wall, and mitral isthmus all required reablation. Unfortunately he continued to have poorly controlled rapid AF and underwent AV nodal ablation. The other patient had a completely intact initial lesion set (standard lesion set plus SVC isolation) but an additional right atrial trigger was noted with isoproterenol and successfully ablated. This was complicated by sinus nodal injury necessitating a pacemaker. Among these four patients, only the one requiring the AV nodal ablation has had documented recurrence following the redo ablation (Supplemental Table 1).

### Complications

Eight procedural complications (6.3%) occurred in seven patients (5.4%, Figure 2B, Table 2). These were noteworthy for five cases of cardiac tamponade. One was intraprocedural in a patient undergoing a fourth procedure for repeated and refractory atypical flutters requiring emergency room presentations. PVI was noted on initial mapping but no non-PV foci or inducible arrhythmias were seen on high dose isoproterenol. VOM ethanol, CS isolation, lateral MI ablation, and SVC isolation were performed prior to the notice of an accumulating effusion thought secondary to right atrial perforation. Pericardiocentesis was performed and ablation of the CTI was deferred. It is noteworthy that he has had good arrhythmia control over 24 months of follow-up except for a single presentation of typical atrial flutter to the emergency room. An additional four patients (3.1%) had a delayed presentation of tamponade (Figure 4), presenting at 12 to 33 days post procedure. One of these four patients had a dissection of the VOM during initial venography and another one of the patients had a possible VOM perforation. Interestingly, two of these patients had unrevealing transthoracic echocardiograms (at three and twelve days post procedure) done for other clinical reasons in the weeks before presenting with tamponade (Figure 4). On a third patient, the hematocrit on the pericardial fluid was assessed due to a dark red appearance and was found to be less than 1%.

**Table 2.** Procedural efficacy and complications.

|  |  |
| --- | --- |
| Patients with rhythm assessment > 90 days follow-up | 94 |
| Follow-up duration (months, range) | 3.0 – 30.1 |
| Median | 7.1 |
| Average | 9.5 |
| Assessment methods |  |
| *Standard electrocardiography only* | 41.5% (39) |
| *Wearable monitor* | 41.5% (39) |
| *CIED* | 17% (16) |
| Recurrence | 14% |
| *After final procedure* | 11% |
| Clinical failure | 5% |
| *After final procedure* | 2% |
| Redo procedure after VOM | 4.3% (4) |
| *Recurrence rate after subsequent redo* | 25% (1)* |
| *Ultimate AV nodal ablation* | 1.1% (1)* |
| Procedurally related complications | 6.3% (8) |
| *Intraprocedural tamponade* | 0.8% (1) |
| *Delayed tamponade* | 3.1% (4) |
| *Sinus injury* | 2.4% (3) |
| *Direct result of VOM ethanol* | 0.8% (1) |
| *Other complications* | 0 |

**Figure 4.**
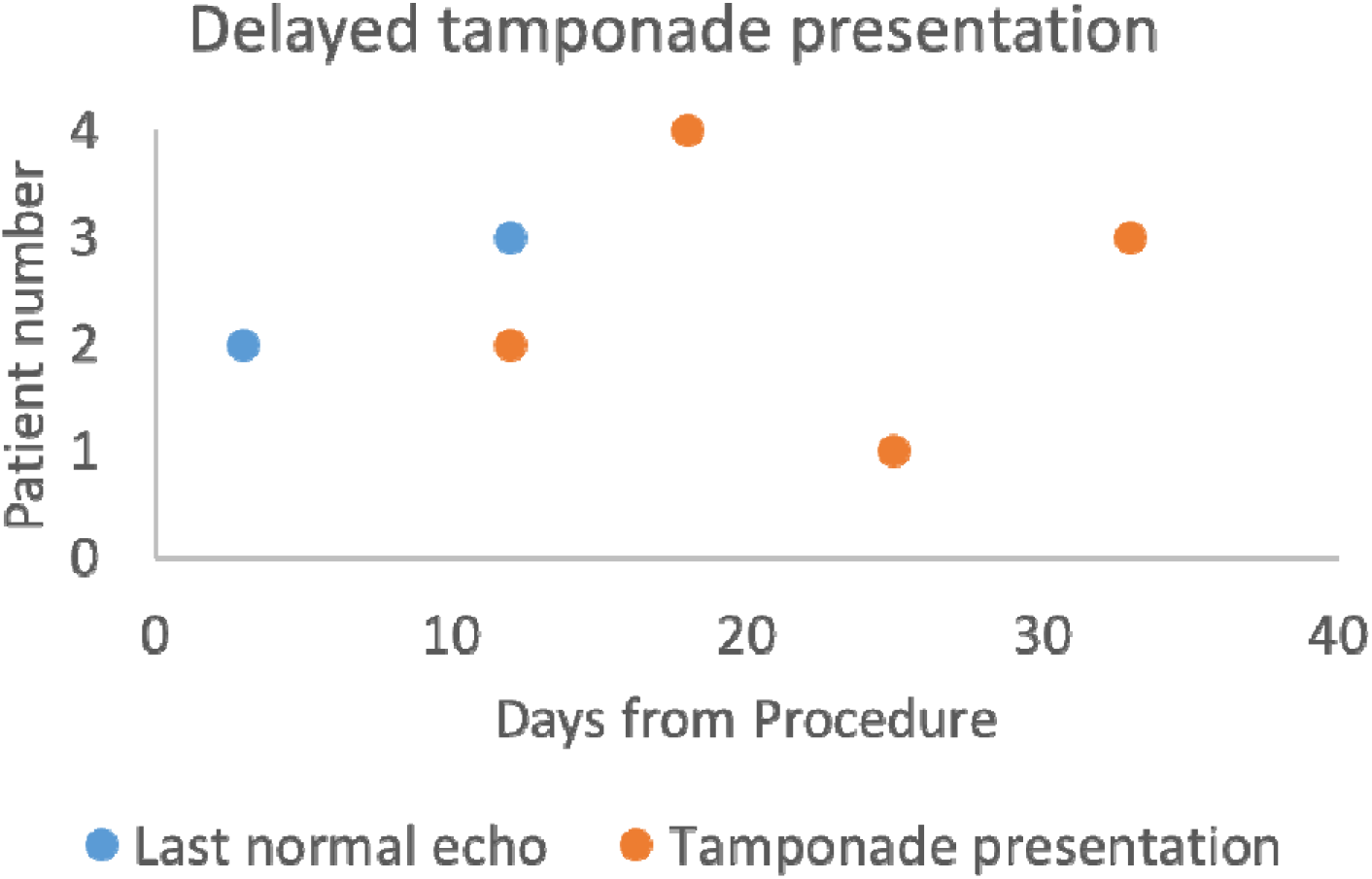
Timing post ablation for four patients with presentations of delayed tamponade. Patients 2 and 3 had normal echocardiograms post ablation in the weeks leading up to presenting with tamponade.

The only other notable complication was sinus nodal injury, occurring in three patients, one of which was also one of the patients presenting with late tamponade above (Patient 3, Figure 4). In one, sinus arrest was noted immediately upon the initial VOM ethanol infusion (Figure 5A). This particular patient presented for a first time ablation for paroxysmal AF. After PVI and PWI were performed, mitral annular and typical flutters were each induced respectfully and attention was diverted to completing both MIL and CTI lines. To facilitate MIL block, a VOM ethanol infusion was planned. VOM venography in that case revealed a small VOM with a typical branching pattern without discernable collaterals to the right atrium/SVC or left atrial roof (Figure 5B, 5C). Some low voltage regions were seen in the right atrium in the sinoatrial region (Figure 5D). The other two sinus nodal injury complications were from direct radiofrequency, both in patients undergoing third AF procedures, one from empiric SVC isolation and the other from ablation of a high cristae AF trigger. All three patients recovered some sinus nodal function but due to pauses or chronotropic incompetence underwent pacemaker implantation in the days to weeks that followed. No additional complications including PV stenosis, stroke, esophageal injury, AV block, left atrial appendage isolation, or anaphylaxis from ethanol infusion were noted. Two deaths occurred which were each judged to be unrelated to the procedure. One patient died 40 days post procedure due to intractable bleeding around rupture of an aortic graft remote from the sites of ablation or trans-septal puncture. The other experienced persistent AF recurrence eleven months post procedure and died the following month of intractable mixed cardiogenic and distributive shock.

**Figure 5.**
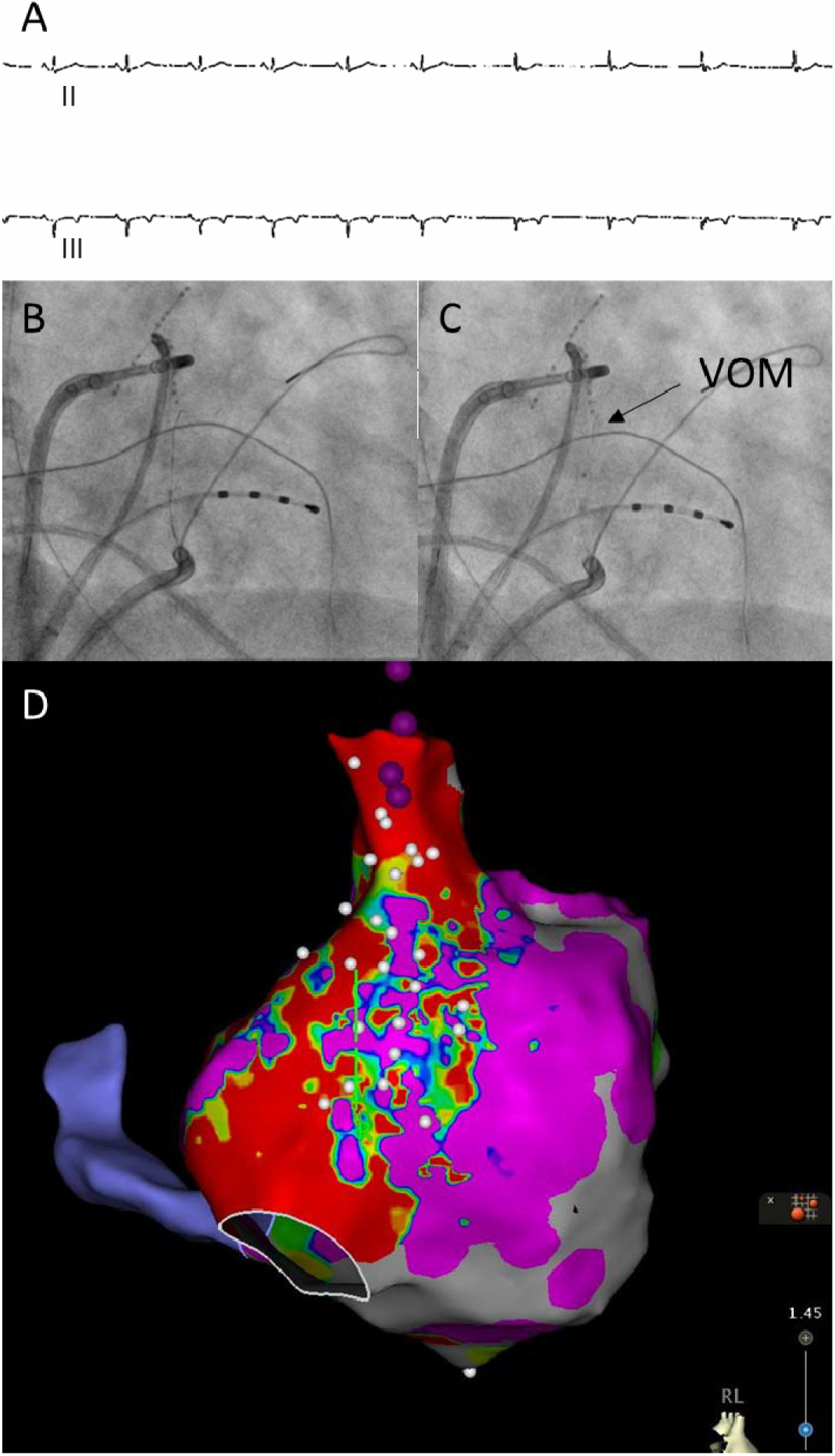
(A) Sinus arrest within 60 seconds of initial EtOH infusion through the VOM. Intracardiac atrial recording was not active during this portion of the procedure as the CS catheter was removed during the VOM infusion. (B) and (C) VOM cannulation and venography. (D) Right atrial low voltage zones on electroanatomical mapping after VOM EtOH infusion.

## Discussion

In this study, we report the feasibility, efficacy, and complication risks related to a moderate volume of VOM ethanol infusions for AF ablation. We demonstrate that VOM ethanol infusion has a learning curve with improved success rates and lowered fluoroscopy times with accumulated experience, even across years of reasonable procedural volume. Rates of VOM perforation, dissection, and overall ethanol infusion success rate were remarkably similar to published reports in a highly experienced center.^8^

A majority of patients had a CIED or wearable monitors to assess arrhythmia recurrence. In the context of a pandemic follow-up system that was often limited to telemedicine without formal rhythm assessments, and the fact that several procedures were performed in the few months leading up to this study, the average follow-up duration was 9.5 months. We acknowledge that with longer follow-up durations and/or continuous rhythm monitoring, more recurrences may have been detected. Nonetheless, we observed extremely low clinical failure and redo procedure rates, and a one year probability of 80% for arrhythmia freedom. This is prominently viewed in light of a significant proportion of long-standing persistent AF patients and the majority of redo procedures enrolled featuring documented permanent PVI. The left atrial size in this study is similar to that of the CONVERGE trial of hybrid ablation,^9^ although the proportion of long-standing persistent AF patients is much lower. Studies of similar success rates in persistent AF patients with less comprehensive ablation approaches have only featured subjects with much smaller atrial sizes.^10^ Unfortunately, the degree that the procedural efficacy is due to the VOM ethanol infusion per say is not possible to assess by this analysis. The standard lesion set in this study encompassed isolation of the posterior wall, CS, mitral isthmus, and cavotricuspid isthmus in almost all cases. Additional ablation beyond this was not insignificant either, particularly in redo procedures. Nonetheless, other studies have questioned the value of these approaches, particularly for ablation lines which were not found to be helpful in a large randomized trial.^3^ Even if the VOM ethanol infusion dominates as the marginal difference accounting for the success in this cohort, the mechanism of benefit remains unclear. In VENUS, the benefit was constrained to the patients with successful mitral isthmus block.^11^ It is possible that the VOM ethanol infusion merely ensures more permanent mitral block^12-14^ or isolation around the left pulmonary vein antra. However additional effects of autonomic denervation^15^, debulking of atrial mass, or direct suppression of ligament of Marshall AF triggers^16^ are plausible as well.

Despite excellent arrhythmia freedom in our cohort, review of these patient outcomes unveiled important safety concerns that also may differ in frequency from those after PVI alone. To the best of our knowledge, we note the first demonstration of sinus nodal injury directly from VOM ethanol infusion. The underlying mechanism is not clear. Although not apparent on venography, we suspect that collateral vessels were present and delivered ethanol to the sinoatrial region. We did not observe any AV block, left atrial appendage isolation, or anaphylaxis that have been described with VOM ethanol.^8^ With respect to the important complication of tamponade we demonstrated a high rate of delayed pericardial effusions. Two of the four patients in this cohort with delayed effusions had unrevealing echocardiograms days after their ablations, only to later present with pericardial tamponade. A third had a hematocrit less than 1% on the pericardial fluid despite a bloody appearance. These findings suggest that the dominant mechanism of delayed tamponade in these cases was inflammatory pericarditis rather than hemorrhagic. Early on in our experience we adjusted our procedural technique to minimize CS related complication. A retention wire is used through the CS guide sheath to avoid CS wall trauma and the IMA catheter is inserted over a wire. Furthermore, we probe for the VOM with a low tip weight angioplasty wire (Suoh wire, Asahi) rather than localize the VOM with contrast puffs. This should lower the risk of VOM perforation/dissection or CS dissection and while we have neared a 100% success rate with VOM ethanol infusion, we have noted a higher rate of delayed tamponade than reported previously in the VENUS study or Bordeaux experience.^8, 10^ The rate is more in line with pericardial effusions in the CONVERGE study of hybrid surgical ablation.^3^ While the low total number of cases leaves open the possibility that our increased rate of delayed tamponade is due to chance alone, interestingly, all four cases of delayed tamponade in our series occurred in the second half of our cohort of patients. We have postulated that as we have become more facile with the procedure, perhaps a brisker workflow with faster ethanol infusions may have contributed. Thus, as discussed in the methods section, while we typically still instill four ethanol injections, we have minimized the total volume of ethanol infused from an average of 10ml to 4-5ml and each injection is instilled slowly over 60 seconds. In addition, in order to avoid contrast induced hydraulic dissections of the VOM, we do not any longer systematically perform VOM venography unless there is uncertainty of the vein identity (non-VOM), the quality of venous occlusion as evidenced by balloon movement, or lack of demonstrable ethanol effects on mapping or ICE. Further study is required to tell if this approach will lower the rate of delayed effusions related to VOM ethanol. Importantly, noting this complication has recalibrated our judgement and patient selection in deciding when to employ the technique.

This study is limited by its modest number of patients, selected population, and the fact that all cases were performed at a single center with a single operator. In addition, as this study was a retrospective descriptive study without randomization, it cannot provide input into the incremental benefit of the VOM ethanol infusion per say in terms of procedural efficacy. Furthermore, follow-up was performed according to the local standard of care and was not systematic with regards to scheduled visits or rhythm assessments. Thus, recurrences may have been under-recognized.

As advances in ablation techniques and energy sources bring us closer to ensuring permanent PVI,^17, 18^ the role for VOM ethanol and other adjunctive techniques for AF ablation may take on greater importance. Currently, there is a paucity of published experience with VOM ethanol infusion and it is rarely performed outside of select centers of excellence. We believe that cases series such as this add important insights to the literature of this technique, informing operators already performing or considering VOM ethanol infusion in the future.

## Conclusion

In summary, we present the experience of a moderate volume of VOM ethanol infusions in routine practice of AF ablation from a single medical center. Our demonstration of the feasibility of growing this program in a previously inexperienced center should allay trepidation to other operators or readers considering adopting this technique into their repertoire. While the high efficacy is laudable, the rate of delayed pericardial effusions should be noted and operators prepared for this complication with early echocardiographic follow-up and for informed consent discussions with patients. Further study of VOM ethanol infusion is needed to determine the marginal benefit and risks across centers.

## Data Availability

Data produced for the purposes of tables and figures and published in this article is available upon reasonable request to the authors. Any additional clinical details are maintained confidential.

## Acknowledgements

The authors would like to acknowledge our colleagues and EP lab staff for their assistance in reviewing this manuscript. We also want to acknowledge Ashley Brooker, Kayla Morrison, John Foley, Jeffrey Peters, and Sheila Robichaud from Biosense Webster for their clinical expertise and assistance with electroanatomical mapping.

## Funding

This work was not supported by any granting agency.

---

**Supplemental Table 1.** Details of recurrences

| Patient number | Type of AF treated | Timing of recurrence (months) | Clinical Failure of 1<sup>st</sup> procedure | Recurrence details |
| --- | --- | --- | --- | --- |
| 1 | PersAF | 14.1 | N | Single episode of pAF < 24 hours |
| 2 | PersAF | 4.4 | N | Recurrence, later controlled on dofetilide until end of follow-up (14.2 months), which had been ineffective prior to ablation |
| 3 | PersAF | 6.0 | N | 3 pAF recurrences on CIED until end of follow-up 13.7 months, longest episode 6 minutes |
| 4 | LSPAF | 6.8 | N | Single 14 hour paroxysmal recurrence on CIED until end of follow-up 13.3 months |
| 5 | PersAF | 10.2 | Y | Recurrent persAF 10.2 months, Died 11.5 months of shock |
| 6 | LSPAF | 3.0 | Y | Recurrent atypical flutter, underwent redo ablation without subsequent recurrence out to 19 months follow-up |
| 7 | PersAF | 5.6 | Y | Recurrent atypical flutter, underwent redo ablation without subsequent recurrence out to 8.7 months follow-up |
| 8 | pAF | 3.0 | Y | Recurrent atrial fibrillation and emergency presentations, underwent redo ablation and subsequently AV nodal ablation |
| 9 | pAF | 3.0 | Y | Recurrent paroxysmal runs of AF, redo ablation of right atrial trigger complicated by sinus nodal injury, no subsequent recurrence for 16.1 months of follow-up |
| 10 | pAF | 7.5 | N | Recurrent typical atrial flutter, single episode out to 24 months follow-up. Preceding history of repeated and refractory emergency presentations prior to ablation. |
| 11 | LSPAF | 5.4 | N | Presented for single episode of SVT cardioverted in emergency room over 18.5 months of follow-up |
| 12 | PersAF | 16.7 | N | Stopped dofetilide shortly before recurrence of persAF, then resumed with subsequent control |
| 13 | PersAF | 7.6 | N | Single 90 second AF recurrence on CIED over 10.9 months of follow-up |

## Notes

*Conflict of interest:* J.L.M. has received speaking fees from Sanofi Aventis. The remaining authors have no relevant financial or non-financial interests to disclose.

*Funding:* The authors did not receive support from any organization for the submitted work.

### Competing Interest Statement

J.L.M. has received speaking fees from Sanofi.

### Funding Statement

This study did not receive any funding.

### Author Declarations

The Maine Medical Center Institutional Review Board gave ethical approval for this work.

## References

1. Haissaguerre M, Jais P, Shah DC, Takahashi A, Hocini M, Quiniou G, Garrigue S, Le Mouroux A, Le Metayer P, Clementy J. Spontaneous initiation of atrial fibrillation by ectopic beats originating in the pulmonary veins. The New England journal of medicine Sep 3 1998;339:659–666.

2. Clarnette JA, Brooks AG, Mahajan R, Elliott AD, Twomey DJ, Pathak RK, Kumar S, Munawar DA, Young GD, Kalman JM, Lau DH, Sanders P. Outcomes of persistent and long-standing persistent atrial fibrillation ablation: a systematic review and meta-analysis. Europace : European pacing, arrhythmias, and cardiac electrophysiology : journal of the working groups on cardiac pacing, arrhythmias, and cardiac cellular electrophysiology of the European Society of Cardiology Nov 1 2018;20:f366–f376.

3. Verma A, Jiang CY, Betts TR, et al. Approaches to catheter ablation for persistent atrial fibrillation. The New England journal of medicine May 7 2015;372:1812–1822.

4. Valderrabano M, Peterson LE, Swarup V, et al. Effect of Catheter Ablation With Vein of Marshall Ethanol Infusion vs Catheter Ablation Alone on Persistent Atrial Fibrillation: The VENUS Randomized Clinical Trial. Jama Oct 27 2020;324:1620–1628.

5. Pambrun T, Denis A, Duchateau J, Sacher F, Hocini M, Jais P, Haissaguerre M, Derval N. MARSHALL bundles elimination, Pulmonary veins isolation and Lines completion for ANatomical ablation of persistent atrial fibrillation: MARSHALL-PLAN case series. Journal of cardiovascular electrophysiology Jan 2019;30:7–15.

6. Derval N, Duchateau J, Denis A, et al. Marshall bundle elimination, Pulmonary vein isolation, and Line completion for ANatomical ablation of persistent atrial fibrillation (Marshall-PLAN): Prospective, single-center study. Heart rhythm Apr 2021;18:529–537.

7. Pathik B, Choudry S, Whang W, D’Avila A, Koruth J, Sofi A, Miller MA, Dukkipati S, Reddy VY. Mitral isthmus ablation: A hierarchical approach guided by electroanatomic correlation. Heart rhythm Apr 2019;16:632–637.

8. Kamakura T, Derval N, Duchateau J, et al. Vein of Marshall Ethanol Infusion: Feasibility, Pitfalls, and Complications in Over 700 Patients. Circulation Arrhythmia and electrophysiology Aug 2021;14:e010001.

9. DeLurgio DB, Crossen KJ, Gill J, et al. Hybrid Convergent Procedure for the Treatment of Persistent and Long-Standing Persistent Atrial Fibrillation: Results of CONVERGE Clinical Trial. Circulation Arrhythmia and electrophysiology Dec 2020;13:e009288.

10. Hussein A, Das M, Riva S, et al. Use of Ablation Index-Guided Ablation Results in High Rates of Durable Pulmonary Vein Isolation and Freedom From Arrhythmia in Persistent Atrial Fibrillation Patients: The PRAISE Study Results. Circulation Arrhythmia and electrophysiology Sep 2018;11:e006576.

11. Lador A, Peterson LE, Swarup V, et al. Determinants of outcome impact of vein of Marshall ethanol infusion when added to catheter ablation of persistent atrial fibrillation: A secondary analysis of the VENUS randomized clinical trial. Heart rhythm Jul 2021;18:1045–1054.

12. Baez-Escudero JL, Morales PF, Dave AS, Sasaridis CM, Kim YH, Okishige K, Valderrabano M. Ethanol infusion in the vein of Marshall facilitates mitral isthmus ablation. Heart rhythm Aug 2012;9:1207–1215.

13. Nakashima T, Pambrun T, Vlachos K, et al. Impact of Vein of Marshall Ethanol Infusion on Mitral Isthmus Block: Efficacy and Durability. Circulation Arrhythmia and electrophysiology Dec 2020;13:e008884.

14. Gillis K, O’Neill L, Wielandts JY, Hilfiker G, Almorad A, Lycke M, El Haddad M, le Polain de Waroux JB, Tavernier R, Duytschaever M, Knecht S. Vein of Marshall Ethanol Infusion as First Step for Mitral Isthmus Linear Ablation. JACC Clinical electrophysiology Mar 2022;8:367–376.

15. Baez-Escudero JL, Keida T, Dave AS, Okishige K, Valderrabano M. Ethanol infusion in the vein of Marshall leads to parasympathetic denervation of the human left atrium: implications for atrial fibrillation. Journal of the American College of Cardiology May 13 2014;63:1892–1901.

16. Dave AS, Baez-Escudero JL, Sasaridis C, Hong TE, Rami T, Valderrabano M. Role of the vein of Marshall in atrial fibrillation recurrences after catheter ablation: therapeutic effect of ethanol infusion. Journal of cardiovascular electrophysiology Jun 2012;23:583–591.

17. De Pooter J, Strisciuglio T, El Haddad M, Wolf M, Phlips T, Vandekerckhove Y, Tavernier R, Knecht S, Duytschaever M. Pulmonary Vein Reconnection No Longer Occurs in the Majority of Patients After a Single Pulmonary Vein Isolation Procedure. JACC Clinical electrophysiology Mar 2019;5:295–305.

18. Reddy VY, Dukkipati SR, Neuzil P, et al. Pulsed Field Ablation of Paroxysmal Atrial Fibrillation: 1-Year Outcomes of IMPULSE, PEFCAT, and PEFCAT II. JACC Clinical electrophysiology May 2021;7:614–627.

